# Causal Relationship and Lipid Metabolite-Mediated Mechanisms Linking Free Testosterone to Abdominal Aortic Calcification: Insights from NHANES and Mendelian Randomization Analyses

**DOI:** 10.1101/2025.08.06.25333175

**Authors:** Renzhi Chen, Chong Liu, Haizhou Pan, Xinyang Li, Hengzhen Wang, Yingdong Sun, Yang Sun, Yang Qiu, Haidi Hu

## Abstract

**Background:** Free testosterone (FT) demonstrates significant associations with cardiovascular diseases (CVDs). However, the precise relationship and underlying mediators linking FT and abdominal aortic calcification (AAC) remain incompletely understood. This study combined data from NHANES cross-sectional studies with Mendelian randomization (MR) and mediation MR analyses to assess the causal relationship between FT and AAC, and further identified lipid metabolites as potential mediators.

**Methods:** Cross-sectional analysis utilized National Health and Nutrition Examination Survey (NHANES) data collected during 2013–2014, encompassing 2,654 individuals. Associations between severe AAC (SAAC) and FT were investigated using weighted multivariable regression, subgroup analyses, and restricted cubic spline (RCS) regression models. Moreover, to verify causal inference, bidirectional two-sample MR analyses were implemented utilizing European ancestry genome-wide association study (GWAS) data. Subsequently, mediation MR analyses were conducted to quantify the contribution of lipid metabolites to the observed associations.

**Results:** A significant inverse association was observed between FT and SAAC; compared to participants in the lowest FT quartile, those in the highest quartile had a 67.9% lower risk (odds ratio [OR] = 0.321, 95% confidence interval [CI]: 0.194– 0.529, P value [P] < 0.001). The nonlinear inverse relationship between FT and SAAC was further confirmed by RCS regression (P for nonlinearity = 0.010). MR and mediation analyses corroborated the causal impact of FT on AAC (inverse variance-weighted [IVW] OR = 0.969, 95% CI: 0.941–0.997, P = 0.033). Four lipid-associated metabolites mediated approximately 36.13% of this causal link, namely mean low-density lipoprotein (LDL) particle diameter, triglycerides to total lipid ratio in large high-density lipoprotein (HDL) particles, total cholesterol to total lipid ratio in chylomicrons and extremely large very low-density lipoprotein (VLDL), and cholesteryl ester to total lipid ratio in chylomicrons and extremely large VLDL.

**Conclusions:** Elevated FT independently associates with a reduced AAC risk and exerts causal effects mediated by lipid metabolic pathways. These findings highlight FT regulation as a potential target for AAC prevention and provide new insights for therapeutic strategies against vascular calcification (VC).

## BACKGROUND

AAC is a specific manifestation of VC, characterized by the pathological deposition of calcium phosphate crystals and lipoproteins in the intima of the abdominal aorta. This phenomenon indicates increased arterial stiffness along with pathological vascular remodeling(1, 2). As a recognized indicator of systemic atherosclerosis, AAC independently predicts subclinical CVDs and future cardiovascular outcomes, and is closely associated with elevated risk for cardiovascular morbidity and mortality from all causes(3–7). Globally, CVDs remain the leading cause of death, and the prevalence of AAC continues to rise significantly with the aging population(8). Given its high prevalence, role in cardiovascular risk stratification, and direct impact on morbidity and mortality, AAC represents a critical public health issue requiring targeted screening and intervention strategies(9).

In males experiencing angina pectoris, reduced testosterone levels negatively correlate with the severity of coronary artery disease (CAD), implicating testosterone deficiency as a potential contributor to CAD risk(10). Prior research has indicated correlations between reduced testosterone levels and various cardiometabolic risk factors such as obesity, insulin resistance, hypertension, and dyslipidemia(11). Circulating testosterone predominantly binds to albumin or sex hormone-binding globulin (SHBG), with free testosterone constituting merely about 1–2% of the total(12). FT represents the bioavailable portion of testosterone and has emerged as an important biomarker for atherosclerosis and other CVDs(13, 14). In men, hypogonadism contributes to endothelial dysfunction, pro-inflammatory states, and adverse lipid profiles, thereby accelerating atherosclerosis(15). In women, oral testosterone supplementation may decrease high-density lipoprotein cholesterol (HDL-C) but potentially reduce triglycerides, both of which represent significant cardiovascular risk factors(16). FT also demonstrates predictive value for cardiovascular events, and lower FT concentrations are associated with adverse cardiovascular profiles, such as diabetes, hypercholesterolemia, and obesity(13, 17). Despite previous studies confirming correlations between FT levels and diverse cardiometabolic conditions, the specific relationship involving FT and AAC remains insufficiently clarified.

Thus, utilizing cross-sectional data from the NHANES 2013–2014 cycle, we systematically analyzed data from 3,654 individuals to elucidate the association between sex hormone levels and cardiovascular risk in both genders. Moreover, MR was employed to infer causality between FT and AAC, while mediation MR analyses explored whether plasma lipid metabolites functioned as intermediaries in this causal pathway.

## METHODS

## 1. Study Design

### 1.1. NHANES Cross-Sectional

#### Study Design

In the initial stage, we performed a cross-sectional analysis based on NHANES data to evaluate the relationship between FT concentrations and SAAC occurrence. Categorical variables were presented as counts and proportions, whereas continuous variables were expressed as mean ± standard deviations (SD). Statistical differences between individuals with and without SAAC were analyzed using chi-square tests for categorical variables and t-tests for continuous variables. Furthermore, multivariable logistic regression models were constructed to determine the relationship between SAAC risk and FT. To explore possible nonlinear relationships between FT and SAAC, RCS regression models were constructed. Furthermore, subgroup analyses complemented by interaction testing were utilized to examine whether demographic or clinical characteristics modified the relationship between FT and SAAC.

### NHANES Variables

#### AAC

AAC severity was evaluated utilizing dual-energy X-ray absorptiometry (DXA) scans performed at the Mobile Examination Center (MEC) with a sophisticated Hologic QDR 4500A fan-beam X-ray bone densitometer (Hologic Inc.). Calcification severity was quantified according to the Kauppila scoring system (0–24 points), where the abdominal aorta adjacent to the L1–L4 vertebrae was segmented into eight sections, each assigned a visual calcification grade: 0 (no calcification), 1 (<1/3 calcified), 2 (1/3–2/3 calcified), or 3 (>2/3 calcified). Based on clinical thresholds, SAAC was defined as an AAC score greater than 6. Participants were categorized into non-SAAC (AAC ≤ 6) and SAAC (AAC > 6) groups.

#### FT

FT levels were calculated using the validated Vermeulen formula, accounting for testosterone binding dynamics with SHBG and albumin(18). The calculation is expressed as follows:

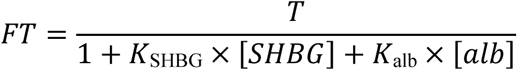

where T represents total testosterone (nmol/L), *K*_SHBG_ denotes the SHBG association constant (1.0 × 10^9^ L/mol), *K*_alb_ denotes the albumin association constant (3.6 × 10^4^ L/mol), [SHBG] represents SHBG concentration (nmol/L), and [alb] is albumin concentration (g/L). Biochemical measurements of total testosterone, albumin, and SHBG were performed on serum samples obtained by trained phlebotomists at the MEC, following standardized NHANES protocols.

#### Covariates

The covariates incorporated into this study included demographic variables, lifestyle behaviors such as smoking and alcohol intake, clinical comorbidities, and extensive laboratory parameters.

### 1.2. MR and Mediation Analysis

#### Study Design

In the second stage, two-sample MR analyses utilizing publicly available summary-level GWAS data were conducted to determine the causal effect of FT (as primary exposure) on AAC (as the outcome)(19). Furthermore, mediation MR analyses following a two-step method were undertaken to examine the role of lipid metabolites as potential mediators in the causal relationship between FT and AAC(20). The total causal effect comprises direct and indirect effects; mediation proportions were computed as the ratio of indirect effects to total effects, accompanied by their respective 95% Cis(21). All analyses conformed strictly to the fundamental assumptions of MR: (1) instrumental variables (IVs) exhibit strong correlations with the exposure; (2) IVs do not correlate with possible confounding factors that might affect the relationship between exposure and outcome; and (3) IVs influence outcomes exclusively via their impact on exposure. These assumptions were carefully assessed through sensitivity analyses. Furthermore, the present investigation complied with STROBE-MR guidelines for reporting(22). The detailed study design is illustrated in Figure 1.

**Figure 1.**
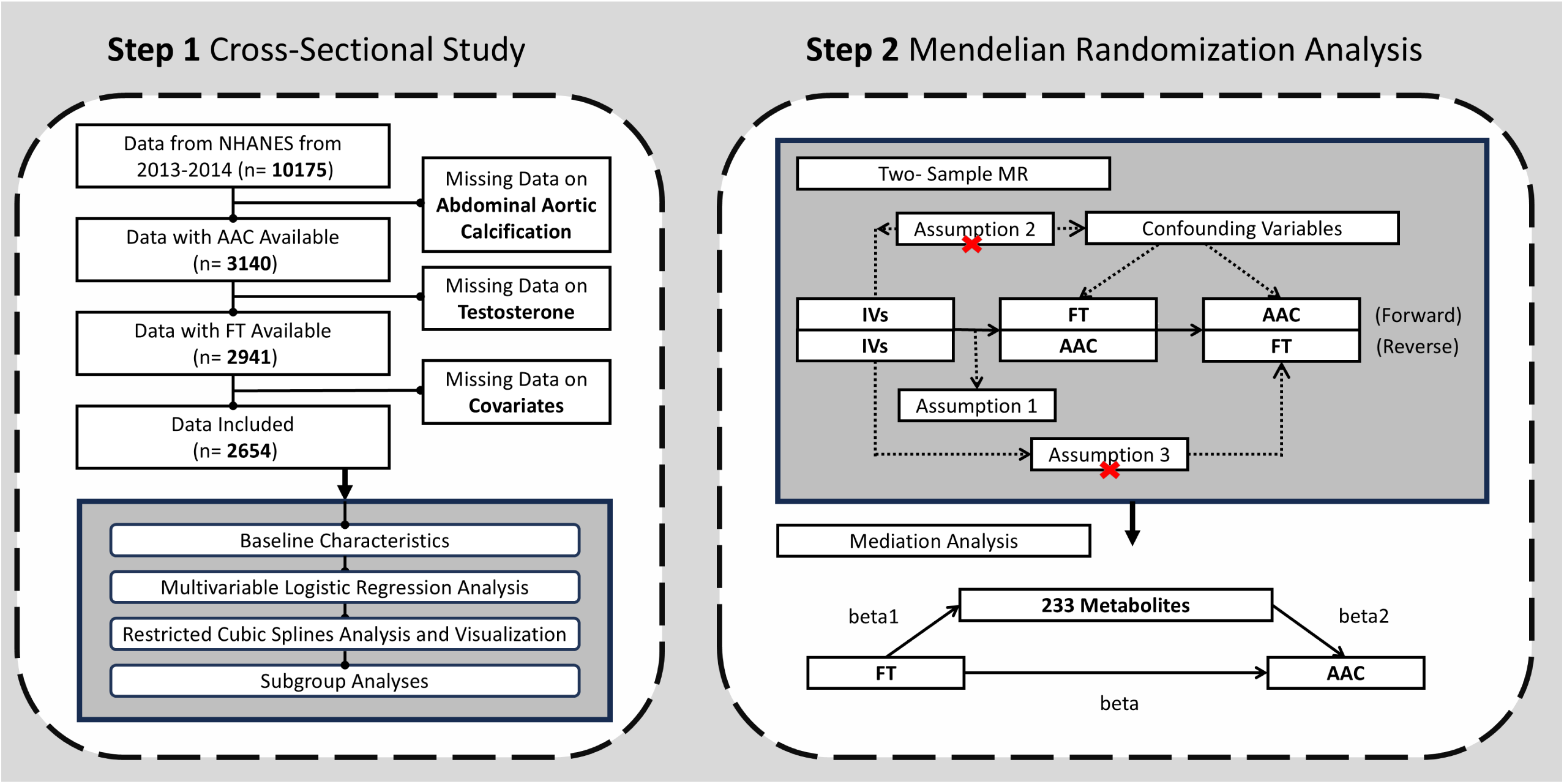
Study design flowchart. NHANES = National Health and Nutrition Examination Survey; IVs: instrument variables; FT: free testosterone; AAC: abdominal aortic calcification.

#### Instrumental Variables

IVs for FT, AAC, and 233 metabolites were selected based on the following criteria: (1) Single nucleotide polymorphisms (SNPs) significantly linked with the exposure (genome-wide significance threshold: p < 5 × 10^-8^); (2) SNPs selection through linkage disequilibrium clumping using the European 1000 Genomes Project Phase 3 reference set (r^2^ < 0.01, window = 250 kb), selecting SNPs with an F-statistic exceeding 10; and (3) addressing palindromic SNPs by harmonizing allelic strands and aligning allele frequencies.

#### Pleiotropy and Heterogeneity Analysis

Cochran’s Q statistic was separately applied in MR-Egger and IVW methods to assess IV heterogeneity related to FT, with P-values greater than 0.05 indicating low heterogeneity. Horizontal pleiotropy was evaluated by MR-Egger intercept and MR-PRESSO global and outlier tests, where P-values above 0.05 implied no significant directional pleiotropy. IVs identified as significant outliers (P < 0.05) were excluded, and MR analyses were subsequently re-performed to confirm the stability of findings.

## 2. Materials

### 2.1. NHANES Data Sources

The National Center for Health Statistics (NCHS) continuously conducts the NHANES, aiming to comprehensively evaluate health and nutritional parameters in the American population(23). Using a multi-stage stratified probability sampling design implemented every two years, NHANES provides nationally representative data(24). For the current analysis, the NHANES 2013–2014 cycle data were initially composed of 10,175 participants. After excluding 7,521 subjects due to incomplete data on AAC, FT, or covariates, a final analytical cohort of 2,654 individuals was obtained.

### 2.2. MR Analyses Data Sources

Summary statistics for AAC were obtained from the GWAS Catalog (GCST90134614) by Sethi et al. (2023), comprising 33,745 individuals of European ancestry (5,594 radiologically confirmed cases and 28,151 controls) with computed tomography-based quantification(25). Genetic associations for FT were derived from the GWAS Catalog (GCST90239825) by Leinonen et al. (2023), including 21,274 participants from the UK Biobank cohort with LC-MS/MS validated serum measurements(26).

GWAS data for the 233 circulating metabolic traits were obtained from Karjalainen et al. (2024)(27). The analysis involving 136,016 individuals identified more than 400 independent genetic loci, nearly two-thirds of which corresponded to genes with likely causal effects. The outcomes revealed genetic associations driven by variations in sample demographics and characteristics, provided insight into genetic determinants of circulating metabolite diversity and their roles in complex disease etiology, and established a valuable resource for further exploration of metabolic-disease linkages.

## 3. Statistical Analysis

R software (version 4.3.2; R Foundation for Statistical Computing, Vienna, Austria) and the “TwoSampleMR” package were employed to perform both MR and cross-sectional analyses. Statistical significance was set at a threshold of p < 0.05.

## RESULTS

### 1. Cross-Sectional Study

#### 1.1. Baseline Characteristics

The analysis included 2,654 individuals with complete AAC measurements (mean age: 58.79 ± 11.96 years; males: 48.2%; females: 51.8%). Among these, 244 participants (9.2%) constituted the SAAC group, which displayed substantial differences in sex hormone profiles relative to controls. Specifically, SAAC-affected men (n = 119) exhibited significantly reduced FT (0.18 ± 0.07 vs. 0.23 ± 0.08 nmol/L; 22% decrease; P < 0.001) and bioavailable testosterone (4.27 ± 1.72 vs. 5.47 ± 1.97 nmol/L; 30% decrease; P < 0.001). In women with SAAC (n = 125), bioavailable testosterone was significantly diminished by 21% (0.15 ± 0.13 vs. 0.19 ± 0.13 nmol/L; P = 0.003), while SHBG levels were notably elevated (81.77 ± 43.65 vs. 72.00 ± 42.88 nmol/L; P = 0.001). Moreover, compared to non-SAAC participants, the SAAC group was significantly older (71.94 ± 8.73 vs. 57.46 ± 11.43 years; P < 0.001), predominantly non-Hispanic White (66.0% vs. 42.2%), and demonstrated higher incidences of hypertension (74.6% vs. 44.5%), diabetes (28.7% vs. 15.1%), smoking history (60.7% vs. 44.8%), and lower total cholesterol levels (4.69 ± 1.05 vs. 5.08 ± 1.13 mmol/L; P < 0.001). Detailed baseline characteristics are presented in Tables 1 and 2.

**Table 1.**
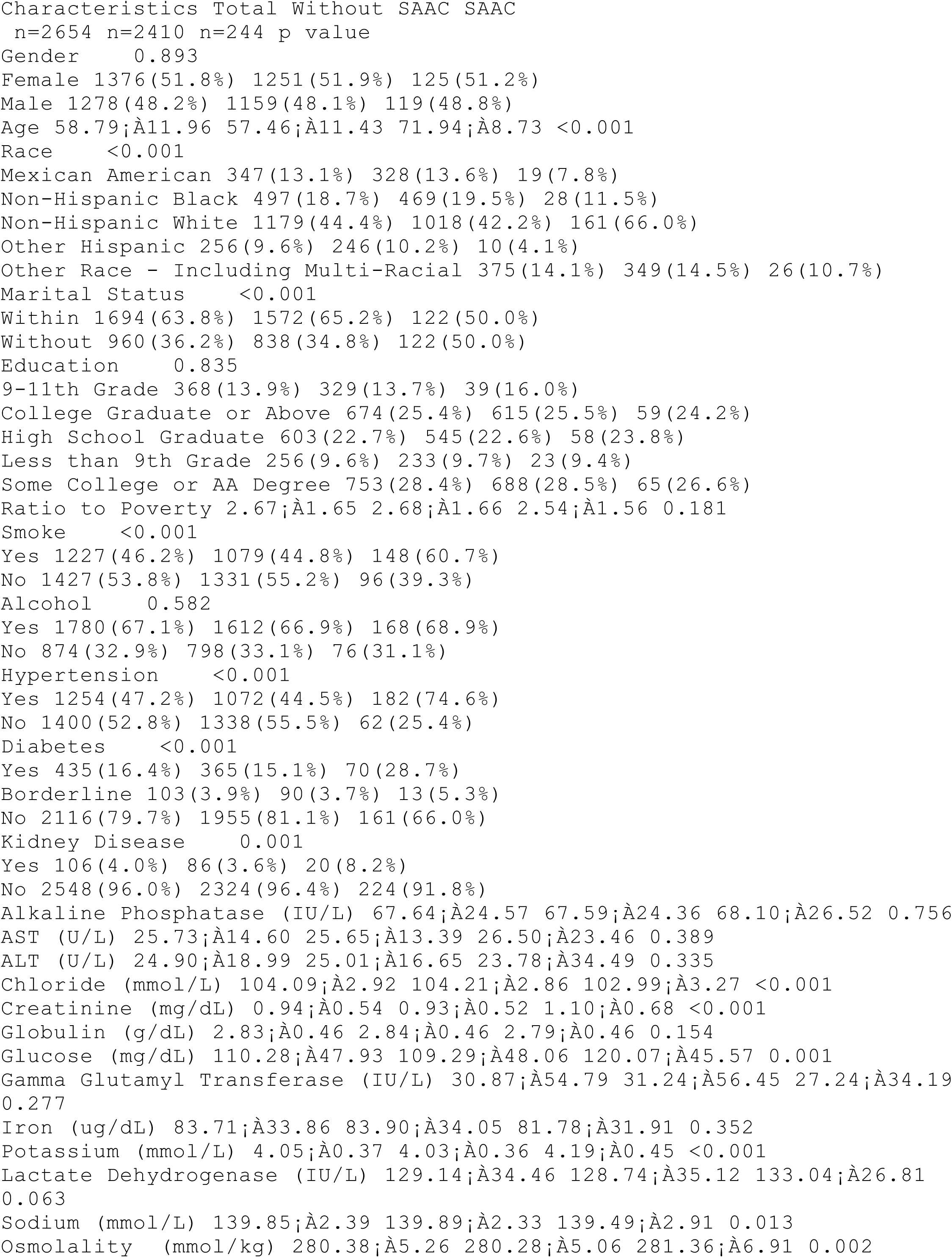
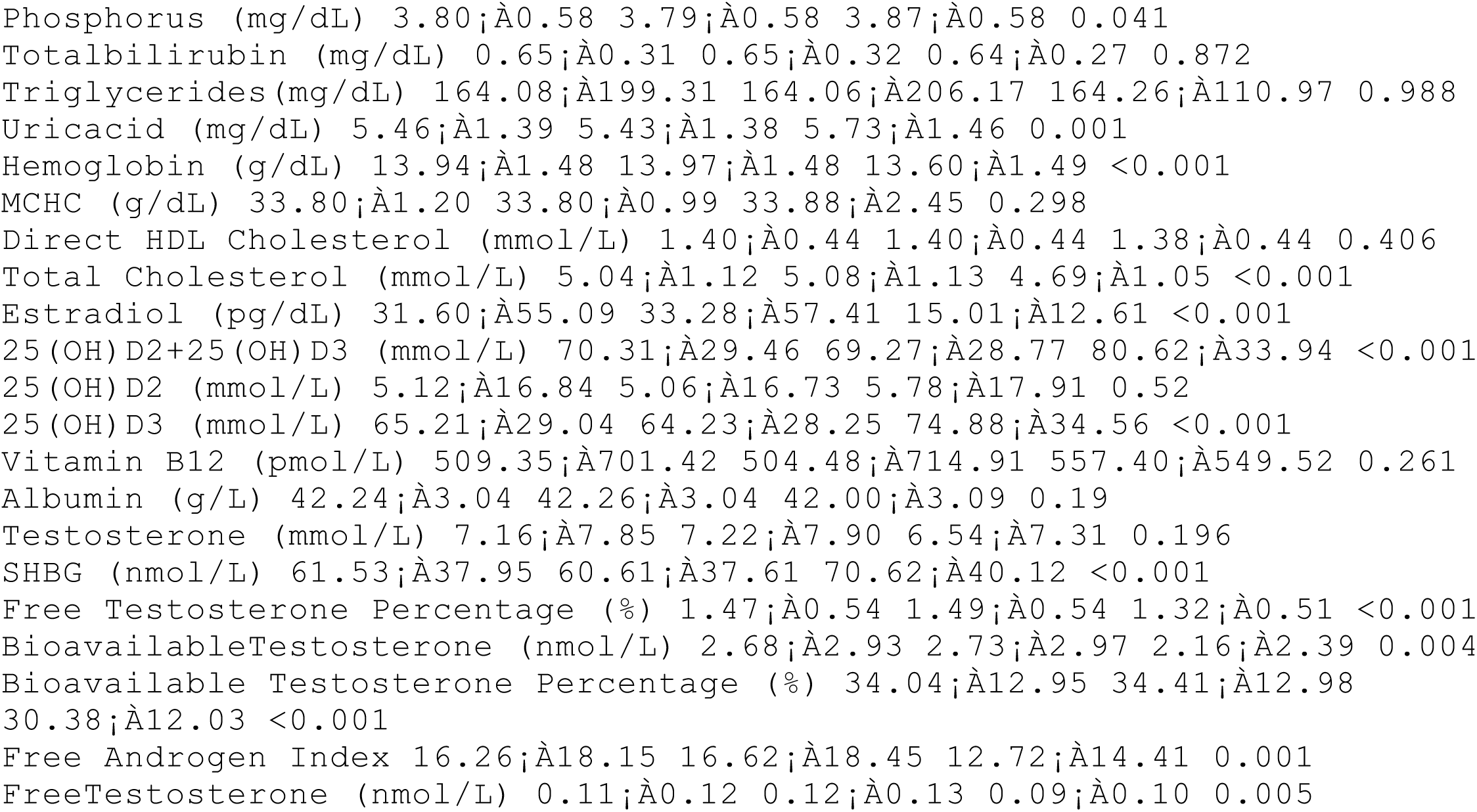
Baseline characteristics of all participants according to SAAC Percentage for categorical variables, mean ± SE for continuous variables. SAAC: severe abdominal aortic calcification; AST: aspartate transaminase; ALT: alanine transaminase; MCHC: mean corpuscular hemoglobin concentration; SHBG: sex hormone binding globulin.

**Table 2.**
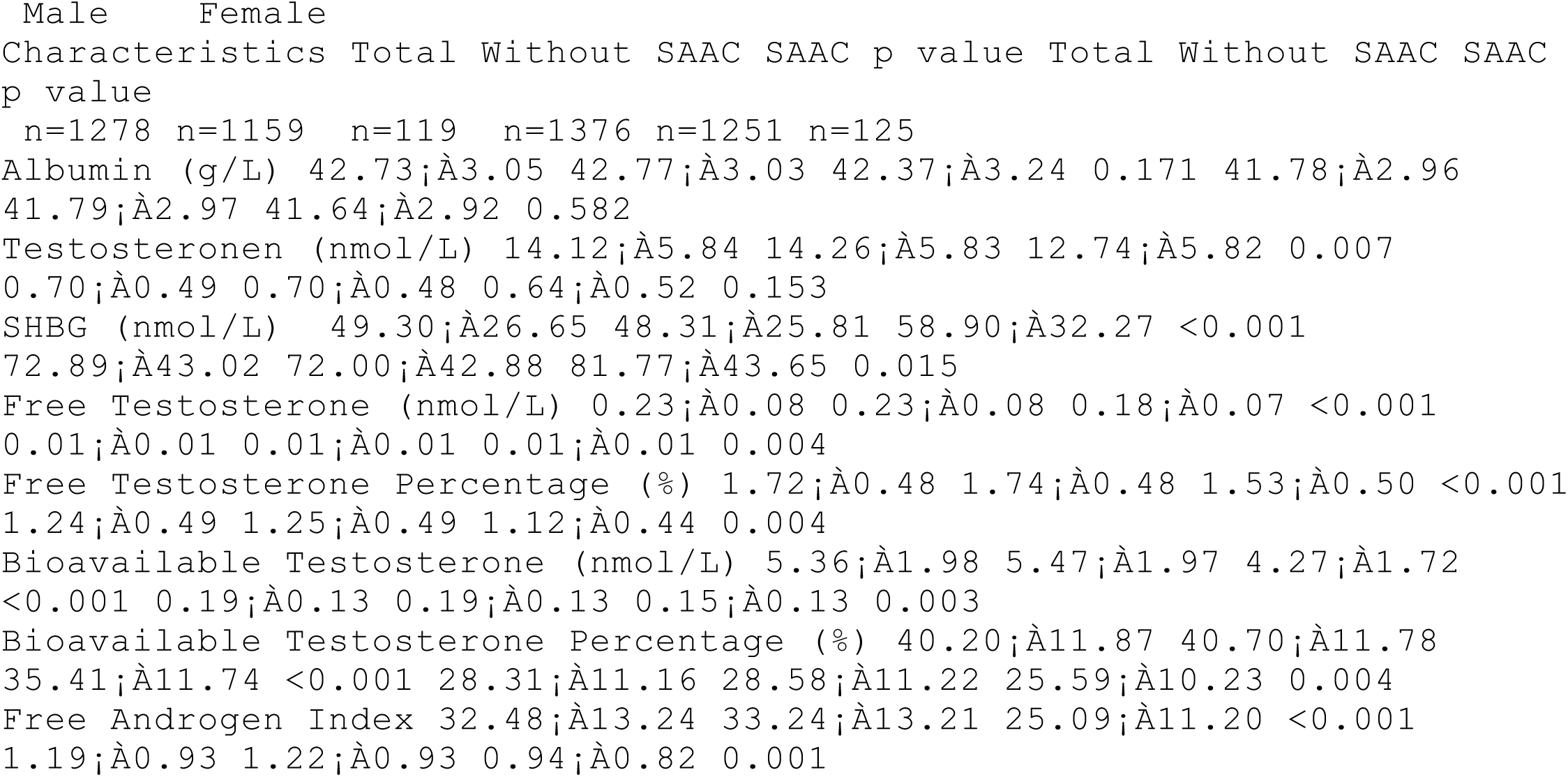
Baseline sex hormone related characteristics of participants according to SAAC and gender Mean ± SE for continuous variables. SAAC: severe abdominal aortic calcification; SHBG: sex hormone binding globulin.

### 1.2. Multivariable Logistic Regression Analysis

An inverse relationship between FT and SAAC risk was consistently observed across all analytical models (Table 3). In Model 1, which involved no adjustments, individuals with FT levels in the top quartile showed a 67.9% lower likelihood of developing SAAC than those in the bottom quartile (OR = 0.321, 95% CI: 0.194– 0.529; P < 0.001). Following partial adjustment (Model 2), this inverse relationship remained significant (OR = 0.247; 95% CI: 0.138–0.439; P = 0.005). After comprehensive adjustment in Model 3, controlling for diabetes, marital status, family income-to-poverty ratio, total cholesterol, chloride, and additional covariates, each incremental rise in FT continued to independently predict a reduced SAAC risk. The inverse relationship between FT and SAAC showed a consistent protective trend across ascending FT quartiles in Model 3 (Q2: OR = 0.477; Q3: OR = 0.497; Q4: OR = 0.394).

**Table 3.**
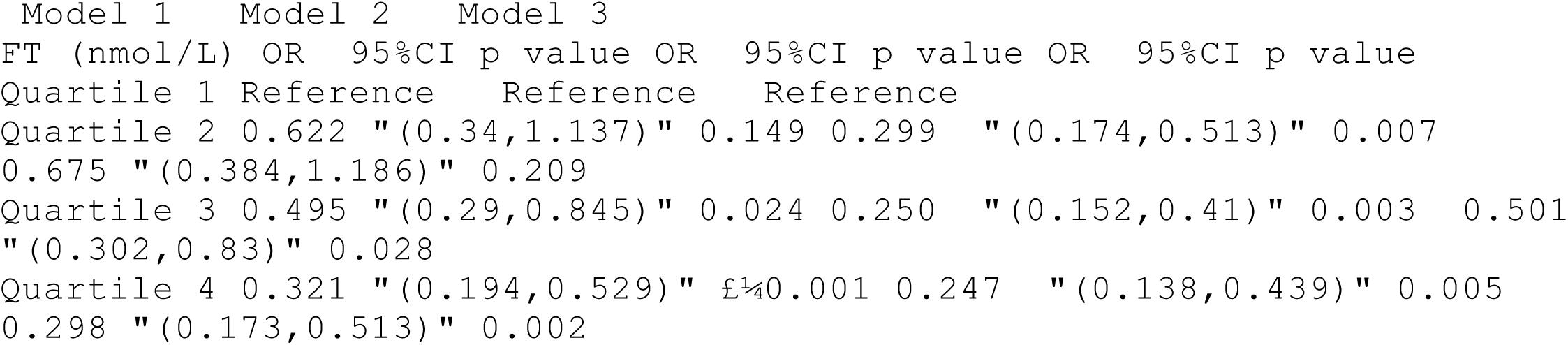
Multivariable regression analysis between FT and SAAC Model 1: Unadjusted model; Model 2: Model adjusted by marital status (in or not in a relationship), family income to poverty ratio and drinking status (at least 12 drinks per year or not); Model 3: Further adjusted by serum total cholesterol (mmol/L), chloride (mmol/L) levels and free testosterone index (FAI). FT: free testosterone.

### 1.3. RCS Regression

RCS regression analysis demonstrated a significant nonlinear relationship between FT concentrations and the risk of SAAC among the entire study cohort (n = 2,654), indicating a gradual reduction in SAAC likelihood with higher FT levels (P for nonlinearity = 0.010). Stratified analyses by sex revealed similar protective patterns at elevated FT concentrations in both males and females. Detailed graphical depictions of the RCS analysis outcomes are provided in Figure 2.

**Figure 2.**
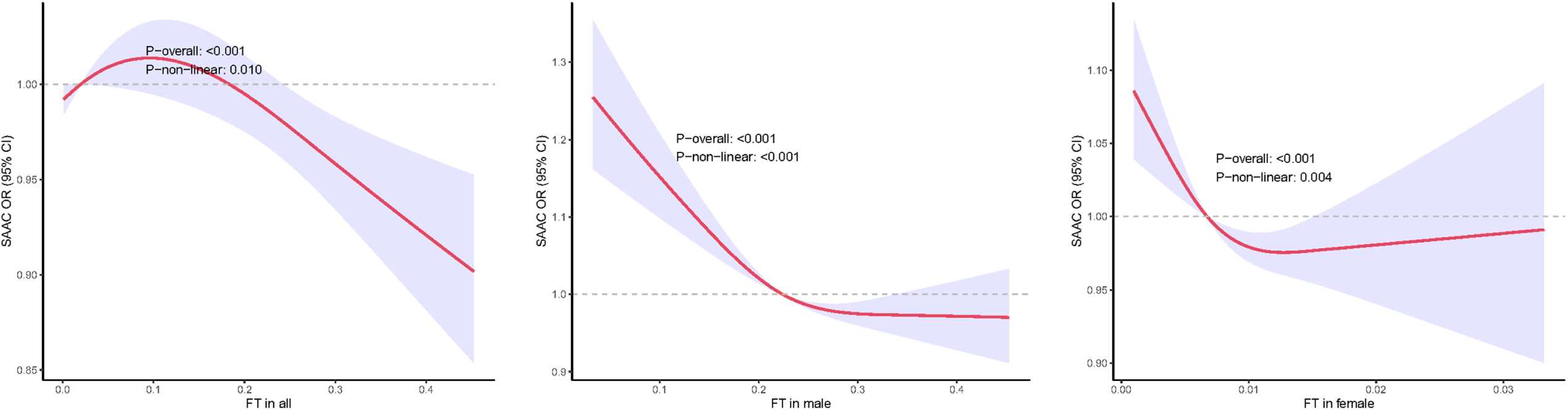
Non-linear association of FT and SAAC from RCS regression The shaded areas represent 95% confidence intervals. SAAC: severe abdominal aortic calcification; FT: free testosterone.

### 1.4. Subgroup Analyses

In subgroup analyses, significant interactions emerged by gender (P interaction < 0.001), smoking status (P interaction = 0.024), alcohol intake (P interaction = 0.028), and presence of hypertension (P interaction = 0.040). Notably, the most substantial protective associations were identified among smokers (OR = 0.863; 95% CI: 0.767– 0.970; P < 0.001) and hypertensive participants (OR = 0.906; 95% CI: 0.856–0.959; P < 0.001). Additionally, non-Hispanic White individuals exhibited meaningful protective effects (OR = 0.924; 95% CI: 0.861–0.991; P = 0.001). Education level was another significant modifier of the association (P interaction = 0.027), revealing the strongest protective impact among those with a college degree (OR = 0.900; 95% CI: 0.833–0.973; P < 0.001), followed closely by participants with partial college education (OR = 0.902; 95% CI: 0.838–0.972; P < 0.001). Comprehensive subgroup analysis outcomes are summarized in Figure 3.

**Figure 3.**
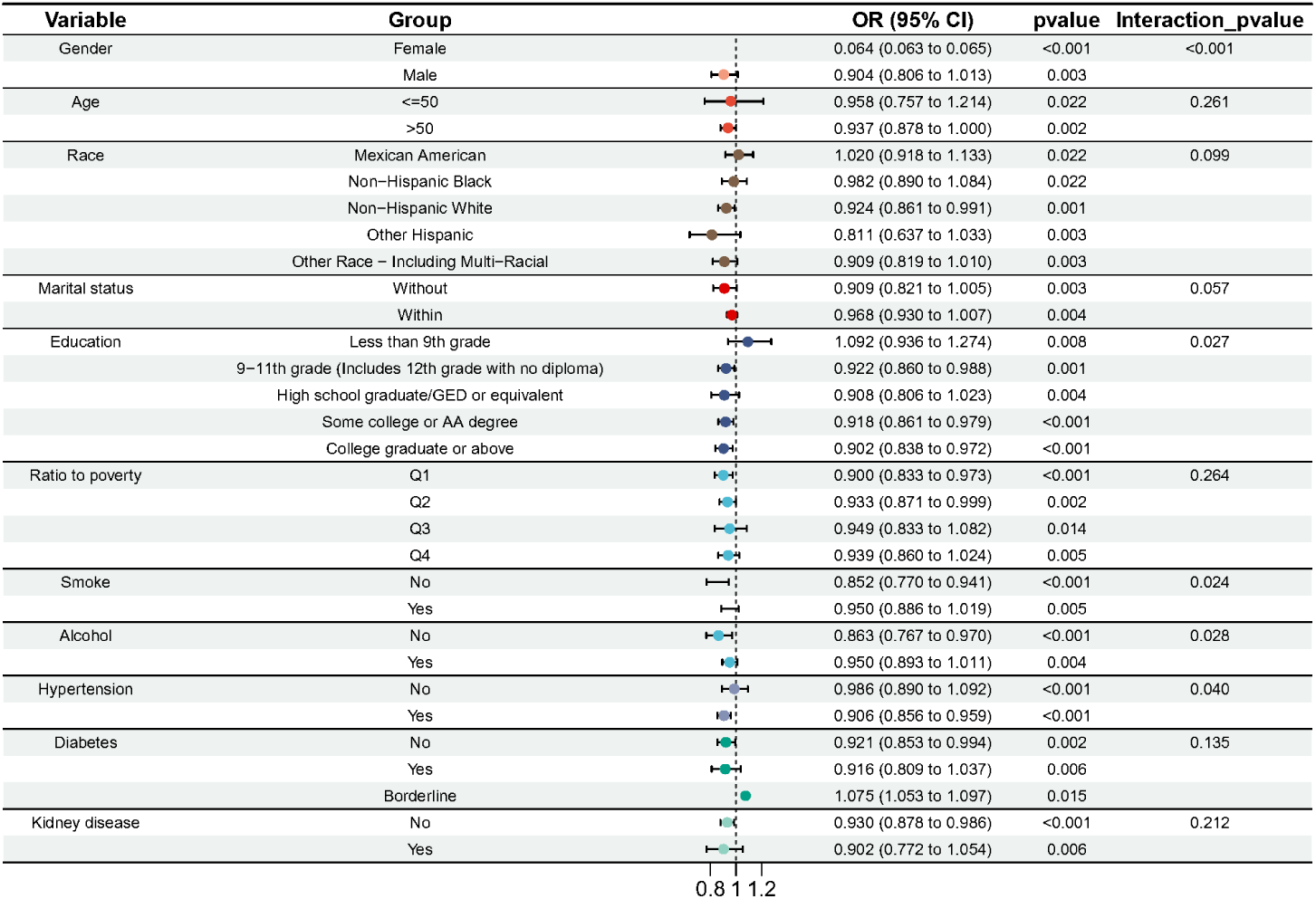
Forest flot to visualize the subgroup analysis results of FT and SAAC Stratification factors were gender, age, race, marital status, education status, family income to poverty ratio, smoke status, drinking status, hypertension, diabetes and kidney disease.

## 2. MR Analysis

### 2.1. Two-Sample MR Analysis

#### 2.1.1. Association between FT and AAC

In MR analysis examining the relationship between genetically predicted FT and AAC, higher FT levels demonstrated a significant protective effect against AAC risk. The primary inverse variance weighted (IVW) method revealed a 3.1% reduction in AAC risk per standard deviation increase in FT (OR = 0.969; 95% CI: 0.941–0.997; P = 0.033). Complementary MR methods yielded directionally consistent yet statistically nonsignificant results: weighted median (OR = 0.973; 95% CI: 0.917– 1.031), MR-Egger (OR = 0.961; 95% CI: 0.910–1.015), and weighted mode (OR = 0.954; 95% CI: 0.882–1.032). Reverse MR analyses examining AAC as a potential causal determinant of FT did not yield statistically significant findings. Detailed outcomes of these two-sample MR evaluations are illustrated in Figure 4.

**Figure 4.**
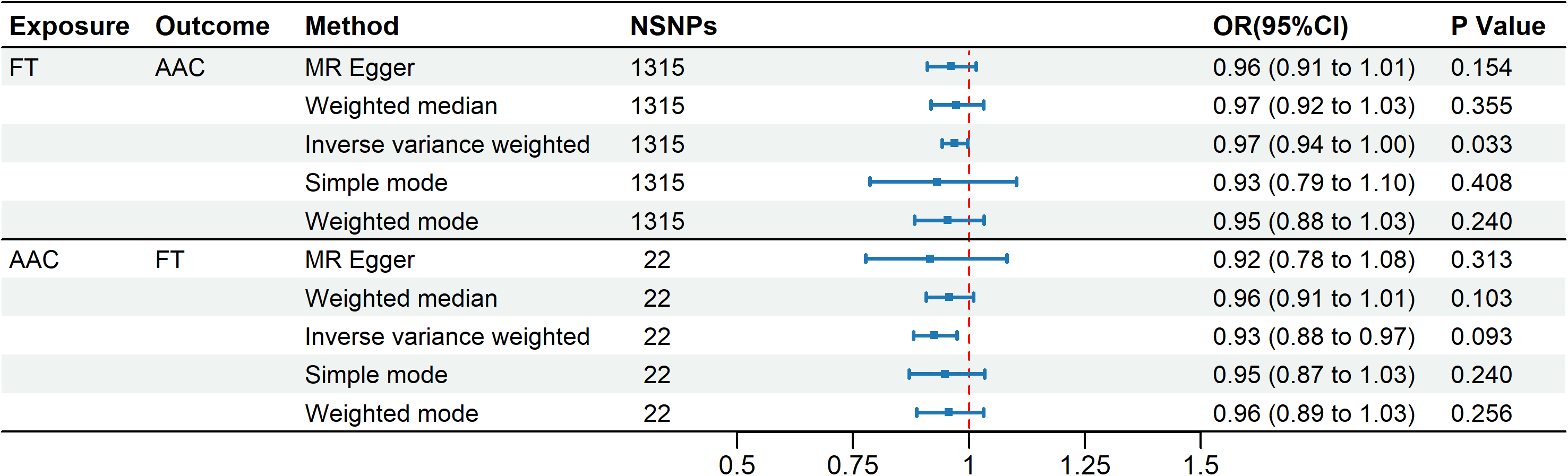
Forest flot to visualize the two-sample MR analysis results of FT and AAC FT: free testosterone; AAC: abdominal aortic calcification.

#### 2.1.2. Association between FT and Plasma Metabolites

Two-sample MR analyses further indicated that genetically instrumented increases in FT were significantly associated with changes in ten plasma metabolites (all IVW P < 0.05). Specifically, higher FT genetically correlated with elevations in creatinine (OR = 1.046; 95% CI: 1.023–1.070; P = 6.71×10^-5^), glycine (OR = 1.028; 95% CI: 1.003–1.054; P = 0.028), acetate (OR = 1.023; 95% CI: 1.005–1.042; P = 0.014), mean LDL particle diameter (OR = 1.035; 95% CI: 1.012–1.058; P = 0.002), the ratio of triglycerides to total lipids within large HDL particles (OR = 1.029; 95% CI: 1.008–1.051; P = 0.007), total cholesterol proportion within chylomicrons and extremely large VLDL particles (OR = 1.031; 95% CI: 1.009–1.054; P = 0.006), and cholesteryl ester proportion in chylomicrons and extremely large VLDL particles (OR = 1.027; 95% CI: 1.004–1.051; P = 0.022). Conversely, FT was significantly associated with reduced levels of phenylalanine (OR = 0.978; 95% CI: 0.957–0.999; P = 0.041), albumin (OR = 0.974; 95% CI: 0.954–0.993; P = 0.009), and tyrosine (OR = 0.977; 95% CI: 0.955–0.999; P = 0.041). Results detailing the associations between FT and metabolites are provided in Table S1.

#### 2.1.3. Association between Plasma Metabolites and AAC

Additionally, comprehensive analyses revealed significant associations between AAC risk and 176 distinct plasma metabolites (all IVW P < 0.05). The identified metabolites spanned various lipid subtypes, including apolipoproteins, lipoprotein particle features, multiple fatty acid categories, and cholesterol transport components across diverse particle sizes. This extensive metabolomic signature underscores the intricate metabolic networks implicated in AAC development. The detailed summary of MR results linking metabolites with AAC risk is available in Table S2.

### 2.2. Two-Step Mediation Analysis

In two-step mediation analyses, four metabolites exhibited significant mediation effects. Regarding mediation analysis, the proportion of the overall testosterone effect explained by mean LDL particle diameter was 7.1% (95% CI: 1.1–25.24%); triglycerides-to-total lipids ratio in large HDL mediated 8.71% (95% CI: 2.64– 26.06%) was mediated by the ratio of triglycerides to total lipids in large HDL particles; the ratio of total cholesterol to total lipids in chylomicrons and extremely large VLDL mediated 7.11% (95% CI: 1.59–30.12%); and the ratio of cholesteryl esters to total lipids in chylomicrons and extremely large VLDL accounted for 13.21% (95% CI: 2.34–49.78%). Collectively, these metabolites accounted for 36.13% of FT’s total causal effect on AAC, emphasizing their substantial role within the biological pathway. Results of the mediation MR analysis are summarized in Figure 5.

**Figure 5.**
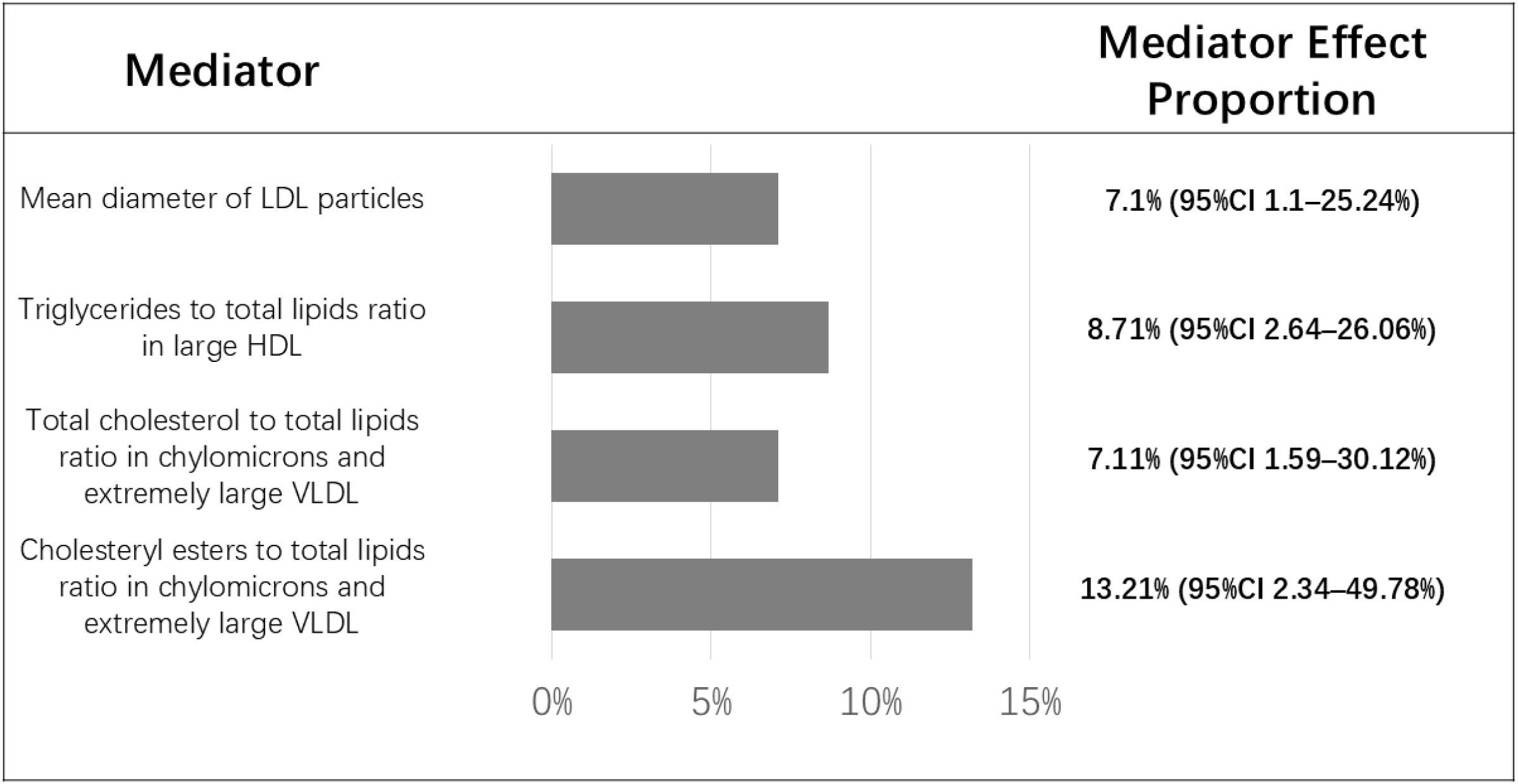
Histograms to visualize the four lipid mediators between FT and AAC by mediation MR

## DISCUSSION

In this cross-sectional analysis based on NHANES 2013–2014 data, we identified a notable inverse association between FT concentrations and the presence of SAAC, suggesting that individuals with elevated FT levels exhibit reduced susceptibility to SAAC. However, observational analyses inherently risk bias from measured and unmeasured confounders, potentially affecting the accuracy of group comparisons. Additionally, such observational approaches cannot conclusively infer causality due to residual confounding and reverse causation possibilities(28). In contrast, MR analyses employ genetic variants as IVs to evaluate causal associations more robustly(29). Our MR findings confirmed a causal relationship between FT and AAC. Additionally, MR mediation analysis further suggested that FT’s beneficial role against AAC was partially attributable to mean LDL particle diameter, triglycerides-to-total lipid ratio within large HDL particles, total cholesterol-to-total lipid ratio, and cholesteryl ester-to-total lipid ratio within chylomicrons and extremely large VLDL.

AAC, a special subtype of VC, has a predictive effect on the prognosis of patients with regional CVDs(30). The prevalence of AAC was the highest among all sites of VC, compared with the calcification of the iliac artery, femoral artery and other arteries(31). Compared to asymptomatic AAAs, AAC score was significantly higher in symptomatic and ruptured AAAs in previous studies, which illustrated the potential that AAC may be a novel tool for AAA rupture risk evaluation(32). A study focusing on AAA tissue specimen had further confirmed the predictive role of AAC on determining AAA rupture risk(33). The size of calcified plaques in normal abdominal aorta was significantly higher in ruptured and symptomatic AAAs than in electively operated AAAs(32). Paradoxically, a radiological study suggested that calcified plaques are smaller in AAA compared to non-aneurysmatic abdominal aortas(34). New et al. demonstrated that small calcified plaques in the AAA cause inflammatory cytokines recruitment which subsequently compromising the structural integrity of the wall, while large calcified plaques are more stable(35). Intimal calcification in abdominal aorta may further aggravate atherosclerosis, thereby increasing risk of arterial occlusive disease like abdominal aortic occlusion(36, 37). Reduction in arterial blood flow caused by AAC mediated through decreased elasticity and increased stiffness may further aggravate arteriosclerosis obliterans (ASO)(38).

Recent studies have highlighted potential associations between sex hormones and CVD risk. Clinical evidence demonstrates that maintaining physiological testosterone levels through supplementation effectively reduces pathophysiological risk factors and clinical severity of CVDs(39). Clinical studies have also reported inverse correlations between FT and carotid intima-media thickness, an early marker of atherosclerosis, independent of age, body weight, and glucose levels(13).Testosterone’s vascular protective mechanisms include dilation of coronary arteries via activation of calcium-activated potassium channels and attenuation of monocyte-endothelial adhesion through downregulation of vascular cell adhesion molecule-1 (VCAM-1), collectively inhibiting atherogenesis(40). Experimental studies further confirm that testosterone relaxes arterial walls via an endothelium-independent mechanism involving potassium efflux(41). Additionally, dihydrotestosterone suppresses atherosclerotic progression by reducing endothelial lectin-type oxidized LDL receptor-1 (LOX-1) expression and inhibiting macrophage-derived intimal foam cell formation(42, 43).

Total and calculated FT levels positively associate with favorable cardiovascular risk factors, including triglycerides, insulin, blood pressure, and HDL-cholesterol. Both cross-sectional and longitudinal analyses indicate that low FT and SHBG concentrations correlate with adverse lipid profiles, characterized by elevated triglycerides and reduced HDL-cholesterol, with lower SHBG levels predicting triglyceride elevation and HDL decline(17). Lipid regulation involves complex metabolic pathways; FT enhances HDL synthesis by stimulating hepatic production of apolipoprotein A1, the primary structural component of HDL particles(44, 45). Concurrently, FT inhibits lipoprotein lipase activity, thereby limiting triglyceride uptake and storage in adipocytes(46). Low FT levels promote visceral fat deposition, leading to increased aromatase activity and accelerated testosterone-to-estradiol conversion, which further suppresses FT. The resulting insulin resistance exacerbates dyslipidemia, characterized by elevated triglycerides and decreased HDL levels(47).

Our mediation analyses indicated that FT may mitigate AAC by modulating four key lipid metabolites. We hypothesize that FT promotes the formation of larger, less atherogenic LDL particles through enhanced hepatic lipase activity, reducing endothelial penetration and cholesterol deposition(45, 48, 49). Simultaneously, FT decreases triglyceride enrichment within large HDL particles, preserving their structural integrity and cholesterol efflux capacity, thus facilitating macrophage-mediated cholesterol clearance(50). Furthermore, FT upregulates lipoprotein lipase and hepatic LDL receptors (LDLR), accelerating hydrolysis and hepatic uptake of triglyceride-rich lipoproteins (TRLs)(51). The mechanism involves decreasing cholesterol and cholesteryl ester concentrations within TRL particles, thereby limiting remnant particle retention and subsequent cholesterol infiltration into the arterial subendothelium. Consequently, the combined effects of enlarged LDL particle size, enhanced HDL function, and diminished TRL cholesterol collectively act to attenuate vascular cholesterol accumulation, ultimately inhibiting VC progression.

Our study has several strengths. The cross-sectional analysis investigated the relationship between FT and SAAC, and the identified causal association was subsequently confirmed through MR analyses. The consistency between MR and cross-sectional findings enhances the reliability and robustness of our results. Moreover, the current study identified particular lipid metabolites as essential mediators, thereby providing insight into the mechanisms by which testosterone modulates atherosclerotic pathogenesis.

Nevertheless, several limitations should be acknowledged. First, a substantial number of participants in the cross-sectional cohort lacked essential AAC or FT measurement data, leading to exclusions that might introduce selection bias. Although NHANES employs sophisticated sampling weights designed to reduce such bias, the exclusion issue persists as a notable concern. Additionally, we focused exclusively on testosterone levels and did not examine potential contributions of other sex hormones to AAC development. Moreover, FT levels vary significantly across gender and age groups, yet we could not perform sex-specific MR analyses due to the absence of sex-stratified AAC data in existing GWAS datasets. Second, MR and mediation analyses assess lifetime causal relationships and mediating effects, rather than relationships at specific time points. Finally, participants in this study predominantly originated from the United States and Europe, limiting the generalizability of our findings to other populations.

## CONCLUSION

This study demonstrates an inverse association between FT levels and SAAC, validated by integrated analyses of NHANES data and MR approaches. Cross-sectional findings indicate that elevated FT confers substantial protection against SAAC. MR analyses confirm a causal protective effect of FT on AAC without evidence of reverse causation. Critically, mediation MR analyses identified lipid metabolism pathways as significant mediators, collectively accounting for 36.13% of the protective effect of FT. These findings position FT as a modifiable biomarker for VC and highlight testosterone-regulated lipid remodeling as a key mechanistic pathway. Despite existing limitations, this study provides foundational evidence supporting FT optimization and lipid-targeted interventions for AAC prevention. Future prospective studies are warranted to confirm the clinical applicability of these findings.

## LIST OF ABBREVIATIONS

(FT): Free testosterone
(NHANES): National Health and Nutrition Examination Survey
(CVDs): Cardiovascular diseases
(AAC): Abdominal aortic calcification
(MR): Mendelian randomization
(SAAC): Severe abdominal aortic calcification
(RCS): Restricted cubic spline
(GWAS): Genome-wide association study
[OR]: Odds ratio
[95% CI]: 95% confidence interval
[P]: P value
(LDL): Low-density lipoprotein
(HDL): High-density lipoprotein
(VLDL): Very low-density lipoprotein
(VC): Vascular calcification
(CAD): Coronary artery disease
(SHBG): Sex hormone-binding globulin
(HDL-C): High-density lipoprotein cholesterol
(SD): Standard deviations
(DXA): Dual-energy X-ray absorptiometry
(MEC): Mobile Examination Center
(alb): Albumin
(IVs): Instrumental variables
(SNP): Single nucleotide polymorphism
(NCHS): National Center for Health Statistics
(IVW): Inverse variance weighted
(ASO): Arteriosclerosis obliterans
(VCAM-1): Vascular cell adhesion molecule-1
(LOX-1): Lectin-type oxidized LDL receptor 1
(LDLR): LDL receptors
(TRLs): Triglyceride-rich lipoproteins

## DECLARATIONS

Ethics approval and consent to participate All protocols used in the research were approved by the Ethics Committee at NCHS, with each participant providing informed consent. Publicly available summary statistics from GWAS datasets were utilized in MR analyses. Ethical approval and informed consent procedures had already been fulfilled by the original GWAS studies through their respective institutional review boards; hence, no further ethical approvals were required for the present analysis.

## Consent for publication

All authors approved the manuscript to be published.

## Availability of data and materials

The raw data utilized in the cross-sectional were downloaded from the NHANES (https://www.n.cdc.gov/nchs/nhanes/continuousnhanes/default.aspx?BeginYear=2013), and GWAS data used in MR analyses were obtained from GWAS catalog (https://www.ebi.ac.uk/gwas/studies/GCST90134614, https://www.ebi.ac.uk/gwas/studies/GCST90239825, https://www.ebi.ac.uk/gwas/publications/38448586).

## Authors’ contributions

RZ. Chen and C. Liu contributed to the conception and design of the study. RZ. Chen contributed to statistical analyses. HZ. Pan, YD. Sun, Y. Sun and XY. Li drafted the article. HZ. Wang revised the article. HD. Hu and Y. Qiu obtained funding and supervised the whole process. HD. Hu is the guarantor of this work.

## Acknowledgements

Not applicable.

